# Artificial Scientific Intelligence for Measurement-burden-aware Modelling and Interpretation of Multi-site Bone Mineral Density

**DOI:** 10.64898/2026.08.30.26361665

**Authors:** Shaowen Xiang, Hongyu He, Ziwen Xie, Hing-Yu Cheng, Hengtong Li, Dianbo Liu

**Affiliations:** National University of Singapore, Singapore; Shanghai Jiao Tong University, Shanghai, China

## Abstract

Agentic workflows can coordinate modelling, but balancing predictive performance, measurement burden and reproducibility is unclear. We developed DXA Agent, an agentic workflow for dual-energy X-ray absorptiometry (DXA) outcomes integrating planning, feature-model refinement, tools, provenance and hypothesis-generating interpretation. Models were independently developed and tested in UK Biobank (5,318 participants) and the National Health and Nutrition Examination Survey (NHANES; 3,777 participants), using cost-efficient and no-limit strategies. Across 20 UK Biobank and three NHANES bone mineral density sites, cost-efficient models achieved lower RMSE and higher R² than the best conventional comparator, with median relative RMSE reductions of 10.9% and 9.9%, respectively. Classification was task dependent: UK Biobank osteoporosis averaged AUROC 0.839 and PR-AUC 0.182, whereas NHANES performance was comparable with conventional models. Higher-burden features did not consistently improve prediction. These retrospective, cohort-internal findings position DXA Agent as an inspectable, measurement-burden-aware research workflow requiring independent prospective validation.

## Introduction

Osteoporosis is a common age-related skeletal disorder in which reduced bone strength increases the risk of fragility fracture, disability and loss of independence. Fractures account for a substantial and growing global health burden as populations age.^1,2^ Dual-energy X-ray absorptiometry (DXA) provides site-specific measurements of bone mineral density (BMD) and remains the reference method used to define osteoporosis from T-scores at clinically relevant skeletal sites according to the 2023 ISCD Adult Official Positions (https://iscd.org/official-positions-2023/). However, DXA requires dedicated equipment and trained operators, making population-wide assessment difficult in many care settings. Fracture-risk calculators and targeted testing can help prioritize assessment, but they do not provide the continuous, multi-site measurements available from DXA. Models based on more readily obtainable health information could therefore support research into triage for definitive DXA assessment, provided that predictive performance is considered alongside the burden of acquiring the required measurements.

Existing clinical risk tools and machine-learning models address parts of this problem, but they do not jointly optimize multi-site DXA estimation, predictor burden and the analytical workflow used to construct the model. FRAX, for example, estimates an individual’s probability of major osteoporotic and hip fracture from established risk factors, with or without femoral-neck BMD; it was not designed to reconstruct BMD or T-scores across skeletal sites.^3^ Machine-learning studies have shown that low BMD or osteoporosis can be identified using demographic, anthropometric, clinical and biochemical variables, and that useful predictive information is often present in routinely collected data.^4,5^ Nevertheless, these analyses are generally implemented as fixed pipelines in which the candidate predictors, model families and evaluation procedures are manually configured for a particular cohort and task. Predictor accessibility is also seldom incorporated as an explicit search constraint. Consequently, a compact model based on routine measurements cannot be systematically weighed against a model that depends on laboratory or imaging-derived variables, even when the latter imposes substantially greater measurement burden.

Large-language-model (LLM) agents offer a way to coordinate these interdependent analytical decisions by combining planning and iterative reasoning with executable computational tools. Recent systems have automated components of clinical machine learning, selected and invoked specialized scientific tools, and decomposed research questions across multiple collaborating agents.^6–8^ These developments create the possibility of moving beyond a single fitted model towards an inspectable research workflow that can examine a cohort, formulate an analysis plan, discover features, build and evaluate models, revise unsuccessful strategies and interpret the resulting evidence. For biomedical applications, however, flexible reasoning alone is insufficient: the workflow must preserve domain constraints, separate language-model reasoning from quantitative execution, record the provenance of analytical decisions and expose trade-offs between predictive performance and data-acquisition burden. Whether these capabilities can be integrated coherently for multi-endpoint population-health modelling remains an open methodological question.

Here we developed DXA Agent, a modular research workflow that links research planning, measurement-burden-aware feature discovery, iterative feature-model refinement, evaluation, provenance and hypothesis-generating scientific interpretation (Fig. 1). We applied the workflow independently to UK Biobank and the US National Health and Nutrition Examination Survey (NHANES), using the predictors and skeletal outcomes available within each cohort. For both datasets, the agent constructed cost-efficient models restricted to more readily obtainable variables and no-limit models that permitted higher-burden measurements. We evaluated continuous multi-site BMD prediction and site-specific classification of low BMD and osteoporosis, compared the resulting models with conventional machine-learning baselines, and examined variation across training seeds. We further characterized model reliance on the selected predictors and used a Scientific Insight Agent to propose and test associations that could motivate subsequent investigation. The objective was to determine whether an agentic workflow could organize reproducible, measurement-burden-aware development and interpretation of cohort-specific DXA outcome models, rather than to establish a replacement for DXA or a clinically deployable diagnostic system.

**Figure 1.**
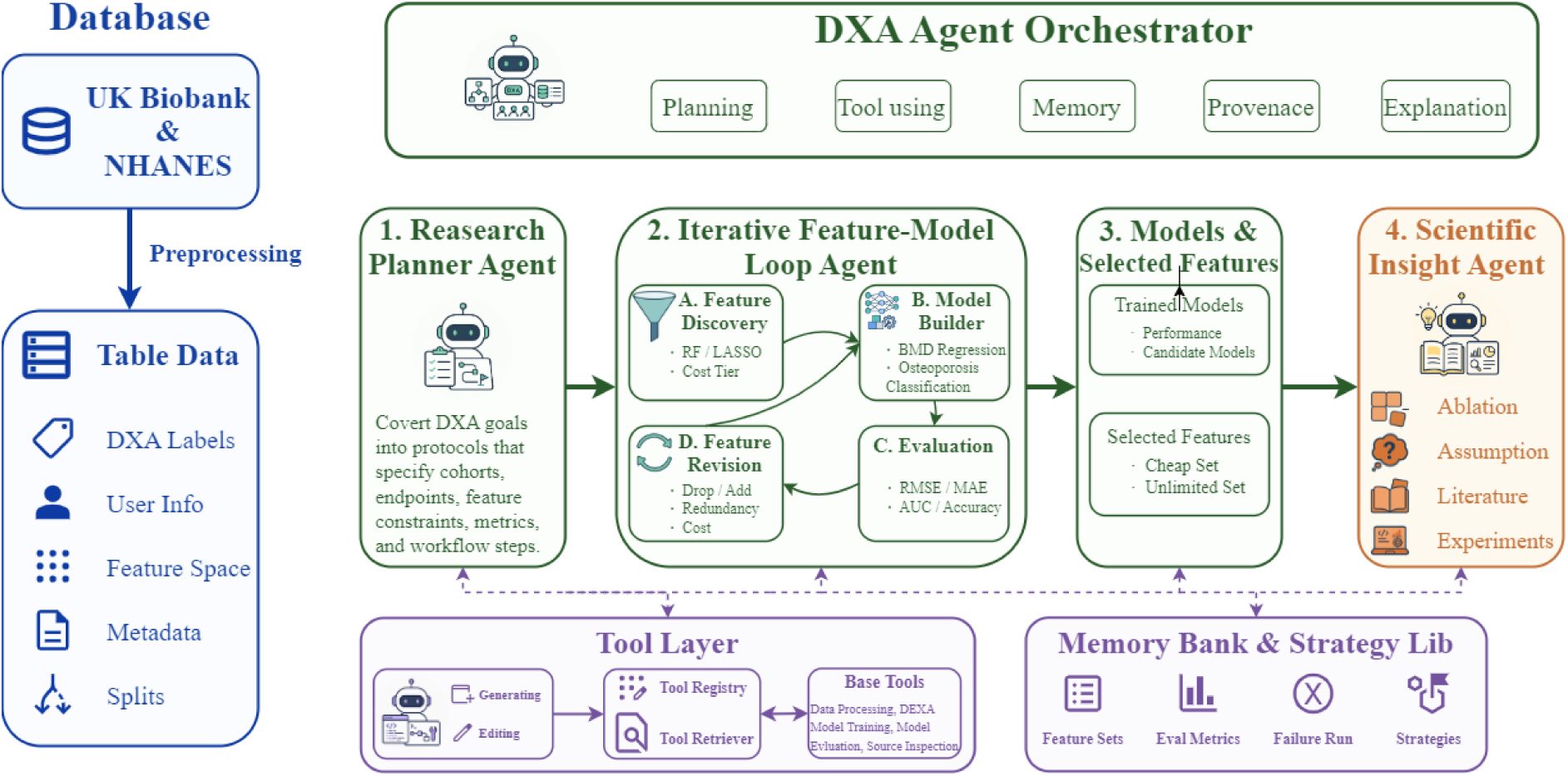
Overall Framework Diagram.

## Results

### Study Cohorts

The analyses included 5,318 UK Biobank participants contributing 5,430 DXA records and 3,777 NHANES participants with linked DXA, survey and laboratory data, as shown in Table 1. The UK Biobank and NHANES cohorts had mean ages of 64.3 (SD 7.5) and 50.3 (SD 15.3) years, respectively. Women comprised 53.7% of the UK Biobank cohort and 47.6% of the NHANES cohort. UK Biobank provided 20 site-specific BMD measurements and six T-scores, whereas NHANES provided three BMD measurements and three corresponding T-scores. Across the available T-score sites, osteoporosis was less common in UK Biobank than in NHANES with site-averaged prevalence, 2.7% versus 8.5%. Models were developed and tested separately within each cohort because their available predictors and skeletal sites differed.

**Table 1.** Baseline characteristics of the UK Biobank and NHANES analytic cohorts.

| Characteristic | UK Biobank | NHANES |
| --- | --- | --- |
| **Cohort and data source** |  |  |
| Data source | UK Biobank DXA and linked phenotypes | NHANES DXA and linked survey and laboratory data |
| Survey cycles | Not applicable | 2013–2014; 2017–March 2020 |
| Participants, n | 5,318 | 3,777 |
| DXA records, n† | 5,430 | 3,777 |
| **Participant characteristics** |  |  |
| Age, years | 64.3 (7.5) | 50.3 (15.3) |
| Female, n (%) | 2,856 (53.7) | 1,797 (47.6) |
| Male, n (%) | 2,462 (46.3) | 1,980 (52.4) |
| BMI, kg m<sup>-2</sup> | 25.7 (3.9) | 27.9 (5.6) |
| **Analysis design** |  |  |
| Candidate predictors, n‡ | 188 | 79 |
| DXA-derived outcomes, n | 26 (20 BMD; 6 T-score) | 6 (3 BMD; 3 T-score) |
| T-score sites, n | 6 | 3 |
| Training set, n | 4,800 | 3,022 |
| Test set, n | 630 | 755 |
| **Bone health distribution** |  |  |
| Normal BMD, % | 67.6 | 49.3 |
| Low BMD, % | 29.7 | 42.2 |
| Osteoporosis, % | 2.7 | 8.5 |

Values are n, n (%), or mean (SD), as indicated. UK Biobank and NHANES were evaluated independently.

Using the workflow shown in Figure 1, the DXA Agent selected a compact, cohort-specific subset of 5-15 predictors under each measurement strategy. From 188 candidate UK Biobank predictors, the final cost-efficient and no-limit configurations each contained eight features as shown in Table 2. The cost-efficient configuration comprised demographic, anthropometric and functional measurements, including age, sex, height, body weight, waist and hip circumference, and left-hand grip strength. The no-limit configuration instead comprised eight brain MRI-derived measurements, including grey-and white-matter volumes and image-registration or image-quality measures. From 79 NHANES candidate predictors, the cost-efficient and no-limit configurations contained 11 and 13 features, respectively. The cost-efficient set emphasized demographic, anthropometric, questionnaire and dietary variables, whereas the no-limit set added laboratory measurements including alkaline phosphatase, calcium, phosphorus, albumin, creatinine and glucose. The resulting feature sets therefore represented distinct measurement-burden profiles rather than nested versions of a common predictor panel.

**Table 2.** DXA Agent selected feature sets.

| **UK Biobank** |  |
| --- | --- |
| **Cost-efficient** | **No limit** |
| Standing height | T1-template mismatch (nonlinear registration) |
| Body weight | T1-template mismatch (linear registration) |
| Age | Peripheral cortical grey matter volume (head-size normalised) |
| Hip circumference | Grey matter volume |
| Height | SWI-T1 image discrepancy |
| Left hand grip strength | Inverted T1 signal-to-noise ratio |
| Sex | dMRI-T1 image discrepancy |
| Waist circumference | White matter volume |
| **NHANES** |  |
| **Cost-efficient** | **No limit** |
| Body mass index (BMI) | Body mass index (BMI) |
| Self-rated general health | Age (years) |
| Age (years) | Standing height |
| Body weight | Serum alkaline phosphatase (ALP) |
| Standing height | Serum total calcium |
| Waist circumference | Serum phosphorus |
| Sex | Serum albumin |
| Race/ethnicity | Serum creatinine |
| Dietary calcium intake | Serum glucose |
| Vitamin D intake | Dietary calcium intake |
| Vigorous work activity | Vitamin D intake |
|  | Dietary protein intake |
|  | Dietary magnesium intake |

Cost-efficient and no-limit feature sets are shown in their selected order for each cohort.

### Automatic DXA Agent

The **DXA Agent** automates end to end development of prediction pipelines for DXA derived bone health outcomes and generates evaluation ready artifacts that support the subsequent performance tables. The agent is run on UK Biobank and NHANES, and it produces dataset specific models that are evaluated on held out test sets. In UK Biobank the agent addresses osteoporosis and low BMD, while in NHANES the agent addresses osteoporosis only. Across runs, inputs are analysis ready DXA outcomes and candidate predictors, and outputs follow a predefined report template that includes the selected feature set, model structure, test set performance, estimated feature prices, and brief rationales for selected features. The agent follows a compact loop that interprets the task, assembles a candidate pipeline, runs preprocessing and quality checks, performs feature selection, trains and evaluates candidate models, then reports and saves artifacts, iterating when checks fail or performance is suboptimal. Outcome definitions, evaluation metrics, and the reporting template are fixed, while model family choice and configuration are searched over, spanning neural network-based models and traditional machine learning models with hyperparameter and structural adjustments.

For each dataset, the agent assembles a consistent analysis matrix, assesses missingness and data quality, and applies rule-based preprocessing. It then composes feature selection strategies using initial screening together with prompt level requirements for novelty and diversity, and it saves the chosen subset and feature importance outputs as reproducible artifacts. Feature costs are estimated by ChatGPT and provided to the agent, which uses these estimates to support cost aware healthcare feature selection and to report an estimated price for the selected feature set alongside a brief reason for inclusion. For each outcome setting the agent outputs two models, a classifier operating on T score derived labels and a regression model for continuous BMD, and it evaluates them using quantitative performance metrics. The agent explores candidates under a small training budget of at most five training runs per task, and it can invoke a Code Agent when additional tools or pipeline adjustments are needed, with the resulting workflow specifications captured in the report artifacts. Full preprocessing rules, feature selection procedures, model families, and tuning details are described in Methods.

### Automatic DXA AI Agent constructs accurate and cost-effective estimation of DXA

Agent generated models were assessed along two tracks: classification of low BMD and osteoporosis using T score derived labels as reference, and regression for continuous BMD prediction. For each task we compared cost-efficient and no-limit DXA Agent strategies to quantify accuracy–resource tradeoffs. UK Biobank analyses covered low BMD, osteoporosis, and multi-site BMD regression; NHANES analyses covered osteoporosis classification and three-site BMD regression. Comparisons to established baselines are included, with definitions in Methods. Comparisons to established baselines are included, with definitions in Methods. Results follow in the next subsections.

### DXA Agent improves continuous BMD prediction across skeletal sites

We first evaluated prediction of continuous BMD on held-out test data, using the mean of three model seeds for each agent configuration as shown in Figure 2a and Table 3. In UK Biobank, the cost-efficient configuration achieved a lower RMSE and a higher R-squared than the best conventional comparator at all 20 BMD sites. Across sites, its median relative reduction in RMSE was 10.9%, and its median gain in R-squared was 0.168. For total-body BMD, the cost-efficient model achieved an RMSE of 0.104 ±0.000 g · cm^−2^ and an R-squared of 0.509 ±0.001, compared with 0.131 and 0.222, respectively, for the best conventional comparator. Performance varied by skeletal site: the cost-efficient model explained more than 60% of the variance in arm, leg and rib BMD, but less than 10% of the variance in head BMD (Supplementary Table S2).

**Table 3.** Regression performance for bone mineral density prediction.

| Model | Metric | a \| UK Biobank |  |  |  |  |  |  |  |  |  |
| --- | --- | --- | --- | --- | --- | --- | --- | --- | --- | --- | --- |
|  |  | Arms | B2-B4 | FN-L | FN-R | FT-L | FT-R | Head | L1-L4 | Legs | Total |
| Linear | RMSE | 0.131 | 0.209 | 0.132 | 0.136 | 0.139 | 0.139 | 0.299 | 0.187 | 0.163 | 0.133 |
|  | R<sup>2</sup> | 0.195 | 0.159 | 0.122 | 0.124 | 0.180 | 0.185 | 0.034 | 0.131 | 0.185 | 0.194 |
| Random forest | RMSE | 0.129 | 0.208 | 0.132 | 0.136 | 0.139 | 0.140 | 0.295 | 0.184 | 0.161 | 0.131 |
|  | R<sup>2</sup> | 0.220 | 0.166 | 0.123 | 0.121 | 0.181 | 0.182 | 0.058 | 0.152 | 0.211 | 0.222 |
| XGBoost | RMSE | 0.128 | 0.205 | 0.133 | 0.136 | 0.138 | 0.139 | 0.298 | 0.185 | 0.160 | 0.131 |
|  | R<sup>2</sup> | 0.238 | 0.190 | 0.119 | 0.114 | 0.183 | 0.185 | 0.041 | 0.149 | 0.213 | 0.218 |
| LightGBM | RMSE | 0.128 | 0.205 | 0.133 | 0.137 | 0.139 | 0.140 | 0.299 | 0.184 | 0.161 | 0.131 |
|  | R<sup>2</sup> | 0.235 | 0.189 | 0.110 | 0.110 | 0.181 | 0.180 | 0.034 | 0.153 | 0.206 | 0.215 |
| Agent (cost-efficient) | RMSE | 0.084±0.000 | 0.176±0.000 | 0.125±0.000 | 0.126±0.000 | 0.124±0.000 | 0.125±0.000 | 0.289±0.000 | 0.172±0.001 | 0.112±0.000 | 0.104±0.000 |
|  | R<sup>2</sup> | 0.674±0.001 | 0.402±0.002 | 0.212±0.001 | 0.240±0.001 | 0.342±0.001 | 0.347±0.002 | 0.094±0.003 | 0.264±0.006 | 0.612±0.003 | 0.509±0.001 |
| Agent (no limit) | RMSE | 0.115±0.000 | 0.209±0.001 | 0.128±0.001 | 0.132±0.001 | 0.134±0.001 | 0.135±0.001 | 0.231±0.001 | 0.178±0.000 | 0.145±0.000 | 0.116±0.000 |
|  | R<sup>2</sup> | 0.380±0.001 | 0.160±0.005 | 0.182±0.009 | 0.172±0.009 | 0.230±0.011 | 0.232±0.007 | 0.424±0.003 | 0.212±0.002 | 0.356±0.003 | 0.390±0.001 |
| Model | Metric | b \| NHANES |  |  |  |  |  |  |  |  |  |
|  |  | Femur neck |  |  |  | Femur total |  |  | Spine |  |  |
| Linear | RMSE | 0.125 |  |  |  | 0.135 |  |  | 0.140 |  |  |
|  | R<sup>2</sup> | 0.332 |  |  |  | 0.291 |  |  | 0.164 |  |  |
| Random forest | RMSE | 0.131 |  |  |  | 0.139 |  |  | 0.144 |  |  |
|  | R<sup>2</sup> | 0.273 |  |  |  | 0.240 |  |  | 0.119 |  |  |
| XGBoost | RMSE | 0.125 |  |  |  | 0.134 |  |  | 0.140 |  |  |
|  | R<sup>2</sup> | 0.331 |  |  |  | 0.295 |  |  | 0.165 |  |  |
| LightGBM | RMSE | 0.127 |  |  |  | 0.135 |  |  | 0.141 |  |  |
|  | R<sup>2</sup> | 0.308 |  |  |  | 0.285 |  |  | 0.156 |  |  |
| Agent (cost-efficient) | RMSE | 0.113±0.000 |  |  |  | 0.114±0.000 |  |  | 0.128±0.001 |  |  |
|  | R<sup>2</sup> | 0.458±0.004 |  |  |  | 0.495±0.004 |  |  | 0.304±0.006 |  |  |
| Agent (no limit) | RMSE | 0.118±0.000 |  |  |  | 0.121±0.001 |  |  | 0.131±0.000 |  |  |
|  | R<sup>2</sup> | 0.406±0.003 |  |  |  | 0.432±0.009 |  |  | 0.265±0.003 |  |  |

DXA Agent values are mean ± SD across three seeds. Conventional comparators are point estimates. Stacking PR-AUC was unavailable and is shown as an em dash.

The UK Biobank no-limit configuration also exceeded the best conventional comparator at most sites, but its gains were smaller and less consistent. It improved both RMSE and R-squared at 18 of 20 BMD sites, with a median relative RMSE reduction of 3.3% and a median R-squared gain of 0.054. At B2-B4 and rib BMD, the no-limit configuration performed less well than the best comparator on both metrics. For total-body BMD, it achieved an RMSE of 0.116 ±0.000 g · cm^−2^ and an R-squared of 0.390 ±0.001. Thus, the inclusion of brain MRI-derived predictors did not improve BMD prediction relative to the lower-burden anthropometric and clinical feature set.

The findings were similar in NHANES. The cost-efficient configuration achieved RMSE values of 0.113 ±0.000, 0.114 ±0.000 and 0.128 ±0.001 g · cm^−2^ for femoral-neck, total-femur and lumbar-spine BMD, with corresponding R-squared values of 0.458 ±0.004, 0.495 ±0.004 and 0.304 ±0.006. These represented improvements over the best conventional comparator at all three sites, with a median relative RMSE reduction of 9.9% and a median R-squared gain of 0.139. The no-limit configuration also improved on the best comparator at all three sites, although its median RMSE reduction (6.2%) and R-squared gain (0.100) were smaller. Site-specific variation across the three seeds was low for both cohorts, with most regression R-squared standard deviations below 0.01 (Supplementary Table S3).

### Detection Low BMD and Osteoporosis in biobank and population survey data

We next evaluated classification of low BMD and osteoporosis using site-specific T-score-derived labels as shown in Figure 2a and Table 4. In UK Biobank, discrimination of low BMD was broadly similar between the two measurement strategies. The site-averaged AUROC was 0.727 for the cost-efficient configuration and 0.722 for the no-limit configuration; the corresponding site-averaged PR-AUC values were 0.568 and 0.575. Relative to the best conventional comparator at each site, the cost-efficient model achieved a higher AUROC at two of six sites and a higher PR-AUC at four of six sites. The largest improvement was observed for total-body low BMD, where the cost-efficient and no-limit configurations improved AUROC by 10.6 and 11.2 percentage points and PR-AUC by 13.1 and 23.4 percentage points, respectively.

**Table 4.**
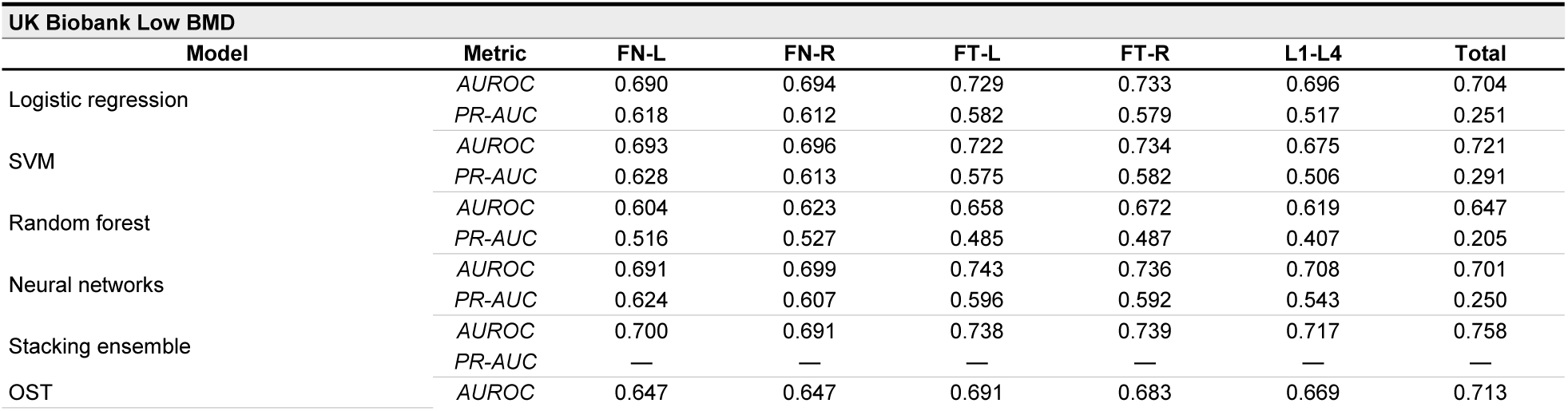
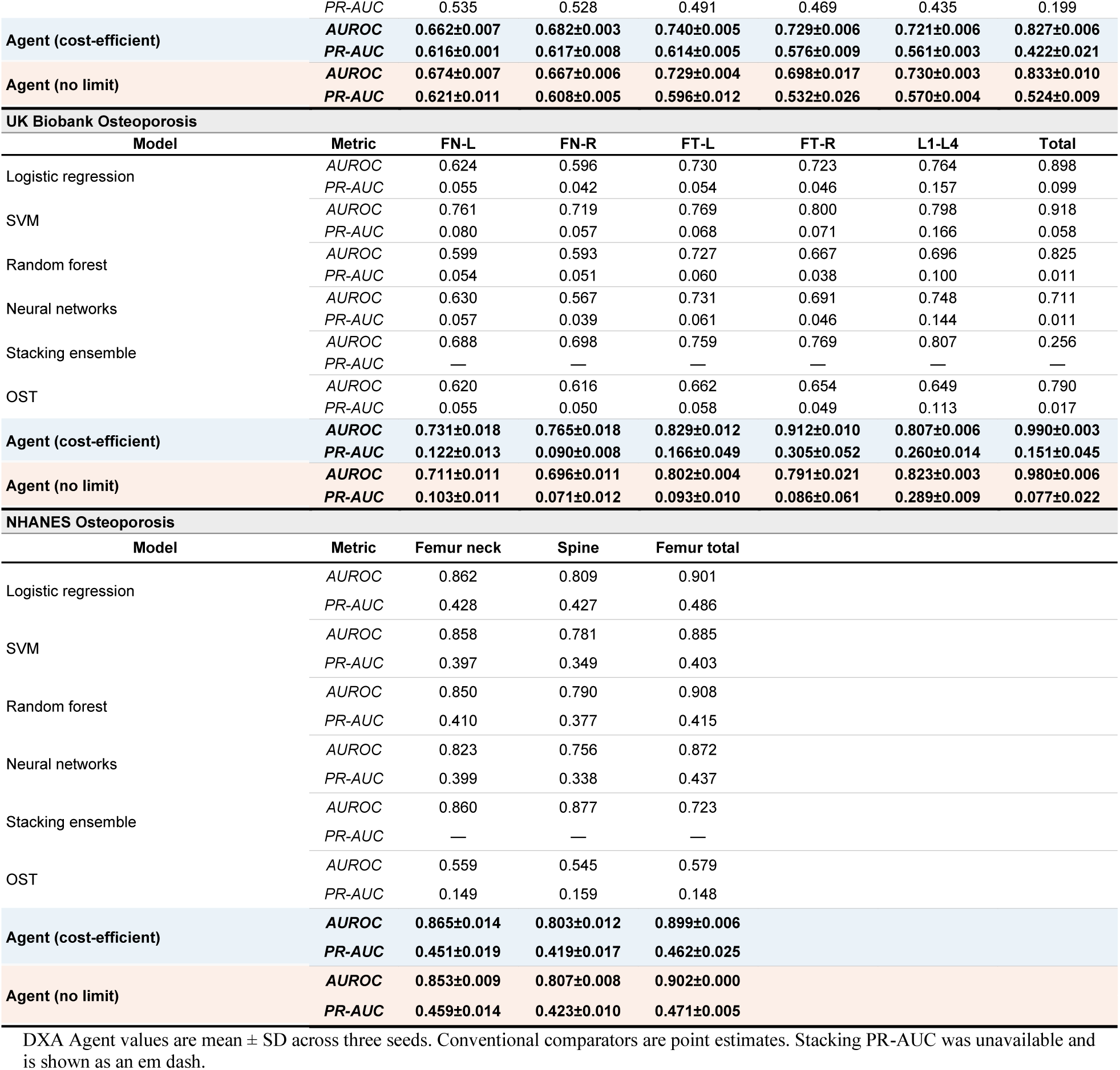
Detection performance for osteoporosis and low bone mineral density classification.

The gains were observed for osteoporosis in UK Biobank. The cost-efficient configuration achieved a site-averaged AUROC of 0.839 and PR-AUC of 0.182, compared with 0.800 and 0.120 for the no-limit configuration. It exceeded the best conventional comparator in AUROC at five of six sites and in PR-AUC at all six sites; the PR-AUC gains ranged from 3.3 to 23.4 percentage points. At the right total femur, for example, the cost-efficient model achieved an AUROC of 0.912 ±0.010 and a PR-AUC of 0.305 ±0.052. Performance nevertheless depended strongly on the metric and site. At total body, an AUROC of 0.990 ±0.003 was accompanied by a PR-AUC of 0.151 ±0.045 and an F1 score of 0.044 ±0.077, illustrating the difficulty of threshold-dependent classification for the rarest osteoporosis endpoints.

In NHANES, agent-only low-BMD performance was similar between strategies: site-averaged AUROC was 0.793 for the cost-efficient configuration and 0.786 for the no-limit configuration, while PR-AUC was 0.799 and 0.800, respectively (Supplementary Table S1). The main comparator analysis focused on osteoporosis. Cost-efficient models achieved AUROCs of 0.865 ± 0.014, 0.803 ± 0.012 and 0.899 ± 0.006 at the femoral neck, lumbar spine and total femur, respectively; the corresponding PR-AUC values were 0.451 ±0.019, 0.419 ±0.017 and 0.462 ±0.025. No-limit performance was similar, with a site-averaged AUROC of 0.854 versus 0.856 for the cost-efficient configuration and a site-averaged PR-AUC of 0.451 versus 0.444. Differences from the best conventional comparator were generally within three percentage points, indicating comparable rather than uniformly superior discrimination in NHANES.

**Figure 2.**
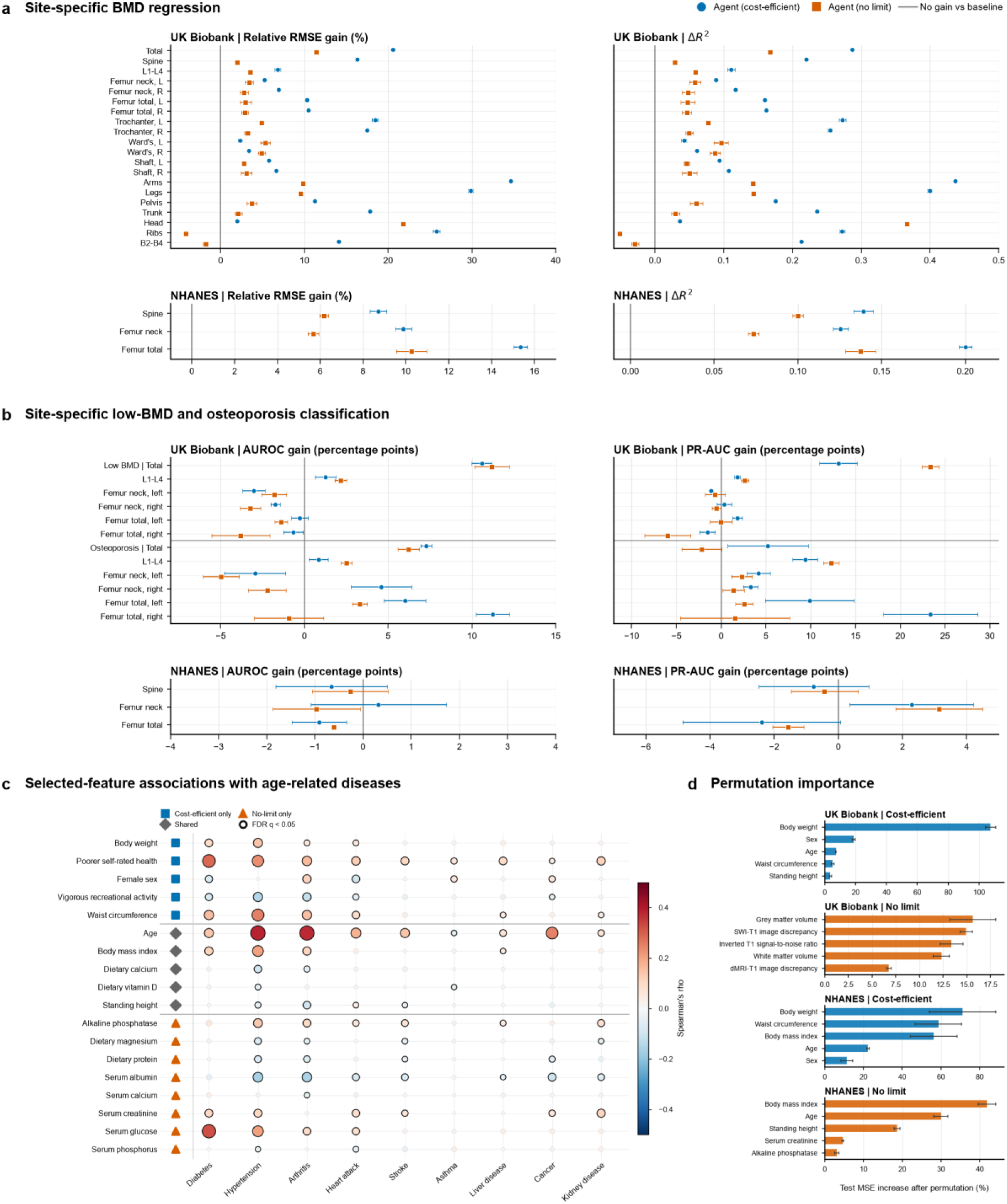
Predictive performance and interpretation of DXA Agent models. a, Site-specific continuous bone mineral density (BMD) regression performance in UK Biobank (top) and NHANES (bottom). Relative root mean squared error (RMSE) gains are shown on the left and changes in R²on the right. b, Site-specific classification performance for low BMD and osteoporosis in UK Biobank (top) and osteoporosis in NHANES (bottom). Differences in area under the receiver operating characteristic curve (AUROC) are shown on the left and differences in area under the precision–recall curve (PR-AUC) on the right. In a and b, circles and squares represent the mean performance across three model seeds for the cost-efficient and no-limit configurations, respectively; horizontal error bars indicate standard deviations. Grey vertical lines indicate no difference from the best conventional baseline selected separately for each endpoint and metric, with positive values favouring the DXA Agent. Relative RMSE gain was calculated as 100 × (baseline RMSE − Agent RMSE) / baseline RMSE. c, Associations between DXA Agent-selected NHANES features and nine age-related disease indicators. Circle colour indicates the direction and magnitude of Spearman’s rank correlation, circle area represents the absolute correlation magnitude, and black outlines indicate associations remaining significant after Benjamini–Hochberg correction across 162 tests (false-discovery-rate q < 0.05). Symbols beside feature names indicate selection by the cost-efficient configuration only, both configurations, or the no-limit configuration only. d, Held-out-test permutation importance for UK Biobank and NHANES multi-output BMD regression models under both measurement configurations. Bars show the five highest-ranked predictors according to the mean percentage increase in standardized multi-output mean squared error after permutation; error bars indicate standard deviations across three model seeds. Correlations and permutation importance quantify association and model reliance, respectively, and should not be interpreted as causal effects.

### Cost-efficient feature sets provide a favourable performance-burden trade-off

Across tasks, greater measurement burden did not translate into a consistent improvement in predictive performance. In UK Biobank, the cost-efficient configuration provided the larger regression gain at nearly every BMD site and performed better on average for osteoporosis classification, whereas low-BMD classification was similar between strategies. In NHANES, the two strategies produced comparable classification results, but the cost-efficient configuration achieved lower RMSE and higher R-squared at each of the three BMD sites. These patterns were observed despite the no-limit configurations incorporating brain MRI-derived measurements in UK Biobank and additional laboratory measurements in NHANES. The comparison was descriptive and does not constitute a formal economic evaluation, but it indicates that routinely obtainable predictors retained most, and in several settings more, of the predictive information captured by the higher-burden feature sets.

### Scientific insights: Model interpretation and associations with age-related diseases

We then examined whether the NHANES features selected for BMD modelling were also associated with nine age-related disease indicators, shown in Figure 2c. Of 162 feature-disease associations, 84 remained significant after Benjamini-Hochberg correction. The largest associations were observed for age with hypertension (Spearman ρ = 0.384) and arthritis (ρ = 0.372), serum glucose with diabetes (ρ = 0.322), poorer self-rated health with diabetes (ρ = 0.288), and waist circumference with hypertension (ρ = 0.253). Cost-efficient features therefore captured broad demographic, anthropometric and self-reported health signals, whereas the no-limit set added disease-aligned laboratory signals, most notably serum glucose for diabetes. These cross-sectional associations were used to characterize the biological and clinical context of the selected predictors and do not establish causal relationships.

Permutation experiments on the held-out test sets showed that the models relied on different predictor domains under the two measurement strategies, shown in Figure 2d. In the UK Biobank cost-efficient regression model, body weight was the dominant predictor: its permutation increased standardized multi-output mean squared error by 107.4% ± 3.2%, followed by sex (18.7% ± 1.0%) and age (7.3% ± 0.2%). The NHANES cost-efficient model similarly emphasized body weight (71.1% ± 17.1%), waist circumference (58.7% ± 12.0%) and body mass index (56.3% ±12.1%). By contrast, the leading no-limit predictors were grey-matter volume and SWI-T1 image discrepancy in UK Biobank, and body mass index, age and standing height in NHANES, with serum creatinine and alkaline phosphatase contributing smaller amounts. These rankings quantify model reliance and should not be interpreted as causal effects, particularly where predictors were correlated.

Targeted leave-one-feature-out experiments provided additional support for selected low-burden predictors in UK Biobank. Removing body weight increased relative regression RMSE by 2.8% and reduced R-squared by 0.036, while removing age increased RMSE by 1.5% and reduced R-squared by 0.019. Removing standing height produced little deterioration in aggregate regression performance. These experiments were limited to the first three selected cost-efficient features and one fitted run per ablation, and therefore served as focused checks of model dependence rather than a complete ranking of feature effects.

The Scientific Insight Agent further generated and tested hypotheses linking selected features to bone health. Within UK Biobank, left-hand grip strength was positively correlated with total-body BMD (Pearson r = 0.526), supporting a hypothesis that muscle function and bone health share mechanical-loading or frailty-related pathways. In the no-limit set, white-and grey-matter volumes were positively correlated with total-body BMD (r = 0.456 and 0.451), motivating a hypothesis of shared ageing-related processes across brain and bone. Table 5 summarizes the agent-proposed pathways, claim strength and representative literature anchors for the selected features. Because these analyses were post hoc and observational, they are presented as hypothesis-generating examples rather than evidence of biological mechanism or clinical utility.

**Table 5.** Scientific insight summary (UK Biobank).

| Strategy | Feature | Primary pathway | Claim strength | Example PMID |
| --- | --- | --- | --- | --- |
| **Cost-efficient** | Sex | Biological differences in bone density | Associational with established mechanism | — |
|  | Body weight | Direct biological pathway | Moderate associational | 42386669 |
|  | Standing height | Direct biological influence | Moderate associational | 42094915 |
|  | Height | Biological pathway | Moderate associational | 42361520 |
|  | Left hand grip strength | Direct biological pathway | Moderate associational | 42380078 |
|  | Waist circumference | Proxy/confounding pathway | Associational with established mechanism | 42382928 |
|  | Hip circumference | Direct biological pathway | Moderate associational | 42382928 |
|  | Age | Biological ageing effects | Associational with established mechanism | 42385930 |
| **No limit** | White matter volume | Direct biological pathway | Moderate associational | 38477810 |
|  | Grey matter volume | Direct biological pathway | Moderate associational | 32625080 |
|  | Inverted T1 signal-to-noise ratio | Direct biological pathway | Moderate associational | — |
|  | T1–template mismatch (linear registration) | Neurological influence on bone density | Hypothesis-generating | — |
|  | T1–template mismatch (nonlinear registration) | Direct biological pathway | Hypothesis-generating | — |
|  | SWI–T1 image discrepancy | Neuroendocrine regulation | Hypothesis-generating | — |
|  | Peripheral cortical grey matter volume (head-size normalised) | Direct biological pathway | Hypothesis-generating | — |
|  | dMRI–T1 image discrepancy | Neurological influence on bone health | Hypothesis-generating | — |

## Discussion

In this study, DXA Agent linked research planning, cost-aware feature discovery, iterative model refinement, evaluation, provenance and scientific interpretation within a single agentic workflow for bone-health modelling. Its most consistent gains were observed for continuous BMD prediction: the cost-efficient configuration outperformed the best conventional comparator at all 20 UK Biobank sites and all three NHANES sites, while using compact sets of routinely obtainable predictors. Classification results were more task dependent. The cost-efficient configuration improved UK Biobank osteoporosis discrimination, particularly PR-AUC for uncommon endpoints, but gains were mixed for low BMD and performance in NHANES was broadly comparable with conventional models. Higher-burden features, including brain MRI measures in UK Biobank and additional laboratory tests in NHANES, did not consistently improve prediction. Together, these findings suggest that the main value of DXA Agent lies not in uniformly maximizing a single performance metric, but in coordinating predictive performance, measurement burden and traceable analytical decisions. The Scientific Insight Agent additionally converted selected features and model behaviour into testable associations, although these outputs remain hypothesis-generating rather than mechanistic evidence. This combination moves the system beyond a fixed prediction model towards an inspectable research workflow, while stopping short of establishing clinical effectiveness.

The predictive findings are broadly consistent with previous machine-learning studies of osteoporosis and low BMD, in which demographic, anthropometric and routinely measured clinical variables carried substantial signal. In the DXA-HIP study, machine-learning models improved discrimination over the Osteoporosis Self-assessment Tool, although the incremental AUROC gains were modest.^4^ An NHANES study using demographic and biochemical variables similarly reported that logistic regression performed at least as well as more complex algorithms, with age, body mass index, sex and routine laboratory measures among the leading predictors.**^错误!未找到引用源。^** These observations help explain why the DXA Agent cost-efficient sets, which emphasized age, sex, body size and physical function, matched or exceeded the no-limit sets. They also caution against assuming that model complexity or acquisition burden necessarily produces clinically meaningful improvement. The selected grip-strength signal agrees with prior NHANES evidence linking greater grip strength to higher femoral-neck and lumbar-spine BMD after adjustment for major covariates.^9^ The observed associations between brain measures and BMD also have biological precedent in genetic and imaging studies, but published findings vary by brain phenotype, skeletal site and population.^10–12^ They should therefore be viewed as prompts for targeted analysis rather than confirmation of a brain-bone mechanism. At the workflow level, recent studies have shown that LLMs can automate parts of clinical machine learning and coordinate domain-specific scientific tasks through planning agents and executable tools.**^错误!未找到引用源。^**^-^**^错误!未找到引用源。^** DXA Agent extends this direction into population bone-health research by making measurement burden an explicit search constraint and by retaining the feature, model, evaluation and interpretation artifacts produced during iteration.

The cost-efficient results have potential clinical relevance because a screening or triage model is useful only if its inputs can be obtained in the intended setting. A compact set based on demographics, anthropometry, questionnaires and physical function could be more practical for prioritizing patients for definitive DXA assessment than a model dependent on MRI or extensive laboratory testing. Such a model should complement, not replace, DXA measurement or clinical judgement, and the present study did not evaluate fracture prediction, treatment decisions or patient outcomes. The comparison between cost-efficient and no-limit strategies was also descriptive rather than a formal health-economic analysis; its implication is a performance-burden trade-off, not demonstrated cost-effectiveness. Methodologically, the separation of LLM reasoning from registered computational tools is important because tool-augmented agents can reduce quantitative errors, but do not eliminate errors in interpretation, tool selection or task specification.^14^ Human review, access controls, executable provenance and predefined validation rules therefore remain necessary safeguards. The stored manifests, feature sets, configurations, predictions and multi-seed summaries provide a basis for reproducibility, consistent with current expectations for transparent reporting of AI prediction studies.^15^ Translation would nevertheless require subgroup analyses across sex, age, ethnicity, socioeconomic position and comorbidity, together with calibration and decision-curve assessment. Lower measurement burden may improve accessibility, but equity cannot be inferred from feature cost alone, particularly when both the source cohort and the availability of imaging or laboratory data can be socially patterned. Prospective evaluation should consequently examine who is referred, who benefits, and whether errors or missing data are concentrated in particular patient groups, in line with guidance for early-stage clinical evaluation of AI systems.^16^

Several aspects of the study constrain interpretation. Both datasets were analysed retrospectively and only within their respective cohorts; independent development in UK Biobank and NHANES demonstrates operation across different data environments, but not transportability of a locked model between populations. The predefined UK Biobank source split contained a small number of participant identifiers in both training and test files, and the earlier UK Biobank regression workflow used a legacy evaluation interface, so these estimates should be treated as exploratory until confirmed using a strictly participant-separated, prospectively locked analysis. UK Biobank also has well-described volunteer and imaging-subsample selection, which may attenuate or otherwise distort population associations.^17^ Some osteoporosis endpoints were uncommon, making AUROC appear favourable even when PR-AUC and threshold-dependent F1 were limited. Variation across three training seeds describes optimization stability, not participant-level uncertainty, and no bootstrap confidence intervals or formal paired tests were available. Feature selection was not repeated as a complete independent agent run for each seed, leaving uncertainty about the stability of the agent’s research decisions. Furthermore, the cost tiers were qualitative and did not incorporate monetary cost, staff time, patient burden or local availability; the cost-efficient and no-limit feature sets were non-nested and sometimes represented different analytic samples, so direct attribution of performance differences to measurement burden is not possible. Permutation importance can redistribute importance among correlated predictors, and the leave-one-feature-out experiment covered only three features in a single fitted run. Finally, the disease correlations and Scientific Insight outputs were post hoc and cross-sectional, the literature retrieval was not a systematic review, and no causal, longitudinal or experimental validation was performed. The system was not evaluated for calibration in new sites, workflow impact, clinician-agent interaction, model update safety or failure rates across repeated end-to-end runs.

Future work should therefore prioritize participant-level external validation with locked preprocessing, feature selection, thresholds and analysis plans, followed by prospective studies of referral decisions and clinical workflow impact. A quantitative feature-cost ontology could incorporate acquisition time, direct cost, invasiveness, missingness and setting-specific availability, allowing the search objective to represent burden more faithfully. Repeated end-to-end runs across LLM backends and data perturbations are needed to assess the stability of planning, feature selection and scientific interpretation, while a predefined agent-error taxonomy should capture unsupported claims, inappropriate tool use, data leakage and failures of provenance. Calibration, decision-curve analysis, subgroup performance and fairness auditing should precede any clinical use. The generated muscle-bone, brain-bone and disease-association hypotheses also require covariate-adjusted, longitudinal and, where appropriate, causal or experimental testing. Subject to these validations, DXA Agent could provide a reproducible and cost-aware research assistant for developing cohort-specific bone-health models and prioritizing follow-up questions. Its present significance is therefore methodological rather than clinical: it shows how an agentic system can organize a complex digital-medicine analysis while preserving explicit constraints, executable evidence and human oversight.

## Methods

### Details of data processing

#### Source datasets and cohort construction

We conducted a retrospective prediction-model study using UK Biobank^18^ and the US National Health and Nutrition Examination Survey (NHANES; https://wwwn.cdc.gov/nchs/nhanes). Cohort-specific models were developed and tested separately in the two datasets using the predictors and skeletal sites available in each dataset.

The UK Biobank analytic files contained 5,430 DXA records from 5,318 participants. The predefined source split included 4,800 training records and 630 test records. Some participants had repeated DXA visits. A retrospective audit identified 30 participant identifiers in both source files, although no identical participant-visit record occurred in both; UK Biobank results are consequently interpreted as exploratory cohort-internal estimates. The NHANES data combined the 2013-2014 and 2017-March 2020 pre-pandemic releases and included 3,777 participants, with 3,022 in the source training set and 755 in the held-out test set. NHANES participant identifiers were unique and did not overlap between sets. Cohort characteristics and analytic sample flow are summarized in Table 1.

#### Outcome definitions and variables

The continuous outcomes comprised DXA-derived bone mineral density (BMD) measurements and corresponding T-scores. BMD, expressed in g · cm^−2^, quantifies bone mineral content relative to the scanned area, whereas a T-score expresses BMD relative to a healthy young-adult reference population in standard-deviation units. UK Biobank provided 20 site-specific BMD measurements and six T-scores, giving 26 continuous regression targets. The six T-score sites were total body, lumbar spine (L1-L4), left and right femoral neck, and left and right total femur. NHANES provided BMD and T-score measurements at the lumbar spine, femoral neck, and total femur, giving six continuous regression targets.

For classification, two binary labels were derived separately at each available T-score site. Low BMD was defined as a T-score ≤ −1.0 and osteoporosis as a T-score ≤ −2.5, consistent with World Health Organization-derived criteria.^19^ Participants meeting the osteoporosis threshold also met the low-BMD definition; these were therefore related binary screening outcomes rather than mutually exclusive classes. This procedure generated 12 classification endpoints in UK Biobank and six in NHANES.

Candidate predictors represented demographic, anthropometric, lifestyle, psychological, cardiovascular, clinical, blood, dietary, and imaging domains. Representative variables included age and sex; height, weight, body mass index, waist and hip circumference, and hand-grip strength; physical activity and self-reported health measures; blood and dietary measurements; and brain MRI-derived measures in UK Biobank. Two measurement strategies were prespecified. The cost-efficient strategy was restricted to routinely obtainable variables, whereas the no-limit strategy permitted higher-burden measurements, including MRI-derived features in UK Biobank and laboratory biomarkers in NHANES.

### Model development

#### Agentic architecture for BMD modelling

DXA Agent was implemented as a modular research workflow that converted a user-defined prediction objective into a sequence of cohort inspection, feature discovery, model training, evaluation, feature revision, and scientific interpretation, shown in Figure 1. A Research Planner Agent specified the cohort, outcome, cost strategy, and workflow steps. Within each cohort, the Feature-Model Loop Agent iteratively selected candidate features, trained prediction models, evaluated performance, and revised the feature set. The system recorded selected features, model configurations, metrics, warnings, and artifact paths in machine-readable files. The reproducible manuscript workflow used the fixed local orchestrator and training tools; an interactive large-language-model interface was not required to reproduce the reported models.

#### LLM backends and code execution

The DXA Agent used a large language model (LLM) as the reasoning and orchestration layer for research planning, tool selection, and interpretation of intermediate results. Computational operations were executed through a registered Tool Layer containing functions for data processing, feature selection, model training, evaluation, and source inspection. Tools accepted structured inputs and returned standardized outputs, allowing the orchestrator to incorporate their results into subsequent decisions.

When an analytical requirement was not covered by the existing registry, a dedicated CodeAgent could generate or edit a Python tool from a natural-language specification. The resulting function was registered in the Tool Layer and made available to the iterative workflow. This separation allowed the orchestrating LLM to focus on experimental reasoning while computational procedures remained explicit and executable. Tool calls, generated or revised code, selected feature sets, evaluation metrics, failed runs, and reusable strategies were recorded in the provenance store and Memory Bank, supporting inspection and reproduction of the workflow. Specific LLM and CodeAgent model versions are reported in the Supplementary Methods to permit updating of the backends without changing the framework definition.

#### Cost-aware feature pricing and task configuration

Candidate predictors were assigned qualitative measurement-burden tiers according to their acquisition modality. Demographic, anthropometric, and routine questionnaire variables were generally treated as low burden, whereas MRI-derived measurements and laboratory biomarkers were treated as higher burden. For each run, the task configuration specified either a cost-efficient strategy, which excluded imaging, genetic, and advanced biomarker variables, or a no-limit strategy, which permitted these measurements. Feature discovery combined random-forest importance^20^ and Lasso-based selection,^21^ merged evidence across methods, applied the strategy-specific constraints, and retained a compact subset for model training.

#### Iterative feature-model search

The DXA Agent was initialized with a baseline multilayer perceptron (MLP) and a registry of feature-selection, training, and evaluation tools. Within each cohort, the Feature-Model Loop Agent then performed four linked steps: feature discovery, model building, evaluation, and feature revision. Feature discovery ranked admissible predictors using statistical evidence and the specified measurement-burden constraint. The selected subset and baseline MLP were passed to the Model Builder, which trained models for the continuous BMD and binary screening outcomes. The Evaluation component returned validation performance at each anatomical site together with class imbalance, convergence, and failure information.

The agent used this feedback to refine both the feature set and the model configuration. It could add or remove predictors, adjust model depth and width, modify dropout and regularization, select a task-appropriate loss function and class-weighting scheme, and revise optimization settings. Candidate changes were required to respond to observed validation behaviour rather than test-set performance. Each revised configuration was returned to the training and evaluation tools, creating a closed research loop in which feature utility, predictive performance, and measurement burden were considered jointly. The loop terminated when the iteration budget was reached or further revision did not improve the prespecified validation objective. The reported workflow used two feature-model iterations, followed by multi-seed refinement of the selected MLP configuration. UK Biobank and NHANES used the same research-loop structure while retaining cohort-specific predictors, outcomes, and trained models.

#### Neural network implementation and validation

The initial prediction model was a multilayer perceptron (MLP) designed for multi-output tabular prediction. During the research loop, the agent refined this template for each cohort, task, and measurement strategy in response to validation feedback. Classification models used class-weighted binary cross-entropy or focal loss to account for endpoint imbalance, whereas regression models minimized multi-output mean squared error. Detailed architectures and optimization settings are provided in the Supplementary Methods.

For model refinement, each source training set was divided into development and validation subsets, with records from the same participant retained in one subset. Preprocessing parameters were estimated from development data and applied unchanged to validation and test data. Model selection, early stopping, and classification thresholds were based on validation performance, and selected configurations were repeated using random seeds.

#### Evaluation metrics

Classification performance was evaluated separately for low BMD and osteoporosis at each anatomical site. Discrimination was assessed using the area under the receiver operating characteristic curve (AUROC) and the area under the precision-recall curve (PR-AUC); PR-AUC was included because several osteoporosis endpoints were uncommon. Threshold-dependent performance was summarized using F1, precision, and recall, and probabilistic error was assessed using the Brier score.

Continuous BMD and T-score predictions were evaluated using root mean squared error, mean absolute error, the coefficient of determination (R), and Spearman rank correlation. Comparisons with conventional machine-learning baselines used matched endpoints and metric definitions. Classification comparators included logistic regression, support-vector machine, random forest, and neural-network baselines; regression comparators included linear regression, random forest, XGBoost, and LightGBM.

Finally, results were summarized across random as the arithmetic mean and sample standard deviation. These summaries quantify variation across training runs and are not participant-level confidence intervals. Comparisons were descriptive, and no formal hypothesis tests or multiplicity-adjusted P values were calculated.

#### Scientific insight analysis

Starting from the selected features, model behaviour, and study objective, the Scientific Insight Agent proposed candidate hypotheses about potential relationships with bone health and translated them into testable analytical questions. It then designed and invoked appropriate data experiments, interpreted the resulting evidence, and revised or prioritized the hypotheses. Experiments could include feature ablation, association and adjustment analyses, subgroup comparisons, interaction tests, and alternative model or feature-set evaluations.

The agent integrated evidence from these experiments with relevant biomedical literature and prior workflow memory to produce structured scientific insights. Each insight linked a hypothesis to its supporting or contradictory evidence, remaining uncertainty, and potential follow-up analyses. This process was intended to support hypothesis generation and research prioritization rather than establish causal mechanisms or clinical conclusions; all generated insights remained subject to expert review and further validation.

#### Statistical analysis and reproducibility

Analyses were performed in Python using PyTorch and scikit-learn. Random seeds were set for Python, NumPy, and PyTorch. Optimized runs stored the feature list, preprocessing objects, model configuration, random seed, validation history, decision thresholds, model checkpoint, endpoint-level metrics, runtime, and participant-level predictions. The unified results manifest provides the canonical link between manuscript values and their source artifacts.

## Data Availability

The UK Biobank data used in this study are available to eligible researchers through the UK Biobank access procedures and were accessed under Application 45925. These data cannot be redistributed by the authors.

https://www.ukbiobank.ac.uk/enable-your-research/apply-for-access

https://wwwn.cdc.gov/nchs/nhanes/Default.aspx

## Data availability

UK Biobank participant-level data are available to eligible researchers through the UK Biobank research application process (https://www.ukbiobank.ac.uk/enable-your-research/apply-for-access) and cannot be redistributed by the authors under the terms of the data-use agreement. The NHANES 2013–2014 and 2017–March 2020 public-use datasets are freely available from the National Center for Health Statistics (https://wwwn.cdc.gov/nchs/nhanes/Default.aspx). Aggregated source data underlying the reported tables and figures will be released with the accompanying code repository.

## Code availability

The source code used to implement the DXA Agent workflow, train and evaluate the models, and generate the reported tables and figures is being prepared for public release. It will be made openly available in a public repository together with environment specifications and instructions for reproducing the analyses. The repository link will be added in a subsequent version of this manuscript.

## Acknowledgments

To do.

## Author contributions

S.X. and D.L. conceived the study. S.X. developed the DXA Agent workflow, curated and analysed the data, conducted the experiments, prepared the figures and drafted the manuscript. H.H. contributed to methodology, model evaluation and interpretation of the results. Z.X. contributed to validation, interpretation of the results and manuscript revision. D.L. supervised the study and contributed to study design and interpretation. All authors critically reviewed and approved the final manuscript.

## Supplementary Information

[Optional: Supplementary Information is provided as a separate PDF file. Supplementary Tables/Data are provided as separate files where applicable.]

